# Reporting standards for outbreak data: A systematic review

**DOI:** 10.1101/2024.05.22.24307752

**Authors:** Vanessa Grégoire, Alex W Zhu, Clint A. Haines, Caitlin M Rivers

**Affiliations:** Johns Hopkins Center for Health Security, Johns Hopkins Bloomberg School of Public Health; Department of Environmental Health and Engineering, Johns Hopkins Bloomberg School of Public Health

**Keywords:** Epidemic, Outbreak, Data Reporting, Reporting Standards, Public Reporting

## Abstract

The current landscape of data reporting for outbreaks is ad hoc and inconsistent. Public health authorities have discretion to determine when, where, how, and what outbreak data to report. This uneven information flow hampers response efforts by decreasing the accountability and transparency needed to build public trust in the public health response. We performed a systematic literature review using the PubMed, EMBASE, MedLine Plus, and Google Scholar databases to identify existing guidelines that address timing, methodology and content of outbreak reporting. Our search strategy produced 46 manuscripts for initial screening to determine eligibility, after which we performed a full-text review of those selected for comprehensive evaluation. We identified four manuscripts that discuss minimum standards and expectations for outbreak reports. Included manuscripts highlight the absence of and the consequent need for minimum standards for what information should be reported to the public during outbreaks. Together, they suggest that the ideal outbreak report should contain information on disease severity, epidemic size and geographic extent, daily and total case count, demographics, transmissibility, signs and symptoms, probable disease transmission and exposure pathways, countermeasure status, and sources of uncertainty. This systematic review of existing guidelines is part of a larger effort to develop consensus guidelines for the public reporting of outbreak data.

## Introduction

A series of significant infectious disease outbreaks have occurred over the past two decades, including the 2002-2003 severe acute respiratory syndrome (SARS)^1^ and 2009-2010 Influenza A (H1N1)^2^ pandemics, the 2012 Middle East Respiratory Syndrome (MERS)^3^ and 2014-2016 Ebola^4^ outbreaks, and the most recent coronavirus disease 2019 (COVID-19) pandemic.^5^ The effectiveness of public health responses to such events depends, in part, on the availability of accurate and up-to-date information about the status and trajectory of an outbreak.^6^ Accordingly, public health departments may choose to issue public situation reports describing recent outbreak developments. Despite its critical importance, the landscape of data reporting for outbreaks is ad hoc and inconsistent.^7,8^

Although many jurisdictions publish regular situation reports to inform the public about the status of an outbreak, each public health authority has discretion in determining what data to include, resulting in significant variations in their content.^7,9^ During an outbreak of the Sudan virus in 2022, for example, Uganda did not consistently report on the number of probable cases,^10–15^ which complicated interpretations of the trajectory of the outbreak and capacity for laboratory confirmation. During an outbreak of meningitis C in Florida in 2022, data reporting was limited to ad hoc updates given to news media.^16,17^ During the COVID-19 pandemic, the manner in which jurisdictions reported test positivity varied, resulting in confusion about how the metric should be interpreted.^18–20^ For example, some jurisdictions excluded multiple tests of the same individual, while others included them regardless of duplication.^19,21,22^

The lack of standardization across outbreaks necessitates that each public health authority independently determines what, when and how to report information. Meanwhile, decision-makers in other jurisdictions who rely on reported information must navigate uneven and varied reporting, complicating their efforts to use data to guide public health responses. This gap has clear consequences. Numerous research studies and reports underscore the importance of having readily accessible public health data.^23,24^ Health agencies rely on publicly available outbreak data to evaluate the threat to their jurisdictions and to guide the development of measures to control the spread of the disease that are proportionate to the risk.^25^ The public relies on media outlets to provide information on the extent of the threat, which allows them to make informed behavioral adjustments.^26^ The inconsistencies in the current data-sharing landscape therefore interferes with the ability to build a shared understanding of the status and trajectory of critical public health events.

To address the gap in outbreak data reporting standards, we undertook a comprehensive effort to establish consensus guidelines for the public reporting of outbreak data. The adoption of consensus guidelines helps to standardize and improve the quality of information reporting in a manner that is easy to understand, replicable, and relevant to decision-makers.^27^ For example, the Consolidating Standards of Reporting Trials (CONSORT) helped improve the quality of clinical trial reporting.^28^

As a first step, we conducted a systematic literature review in accordance with the established protocol for consensus guideline development outlined by the EQUATOR network.^29,30^ The purposes of this review were to identify existing guidelines related to the timing, methods, and content of public reporting during outbreaks and to assess the strengths and limitations of these guidelines with the aim of identifying gaps in the current infrastructure.

## Materials and methods

We invited ten leaders in epidemiology and outbreak response to join the study’s steering committee. Nine experts from the United States Centers for Disease Control and Prevention, the United Kingdom Health Security Agency, the World Health Organization, state public health institutions, and academia agreed to participate. The steering committee members convened a series of virtual meetings to finalize the search strategy and search ontology. The committee adopted the PRISMA^31^ methodology in accordance with best practices for conducting systematic reviews.

### Search strategy

We searched the PubMed, EMBASE, MedLine Plus, and Google Scholar databases using the following search criteria: [Outbreak OR Epidemic OR Disease OR Infectious Disease OR Public Health] AND [Data OR Report] AND [Guidelines OR Framework]. We saved the search results in Covidence for processing. The search strategy produced 46 manuscripts for initial screening.

### Study screening and eligibility determination

We assessed the manuscripts identified through the search strategy using a three-phase review process, as depicted in Figure 1. The three phases comprise an initial screening, a full-text review, and a final analysis.

**Figure 1:**
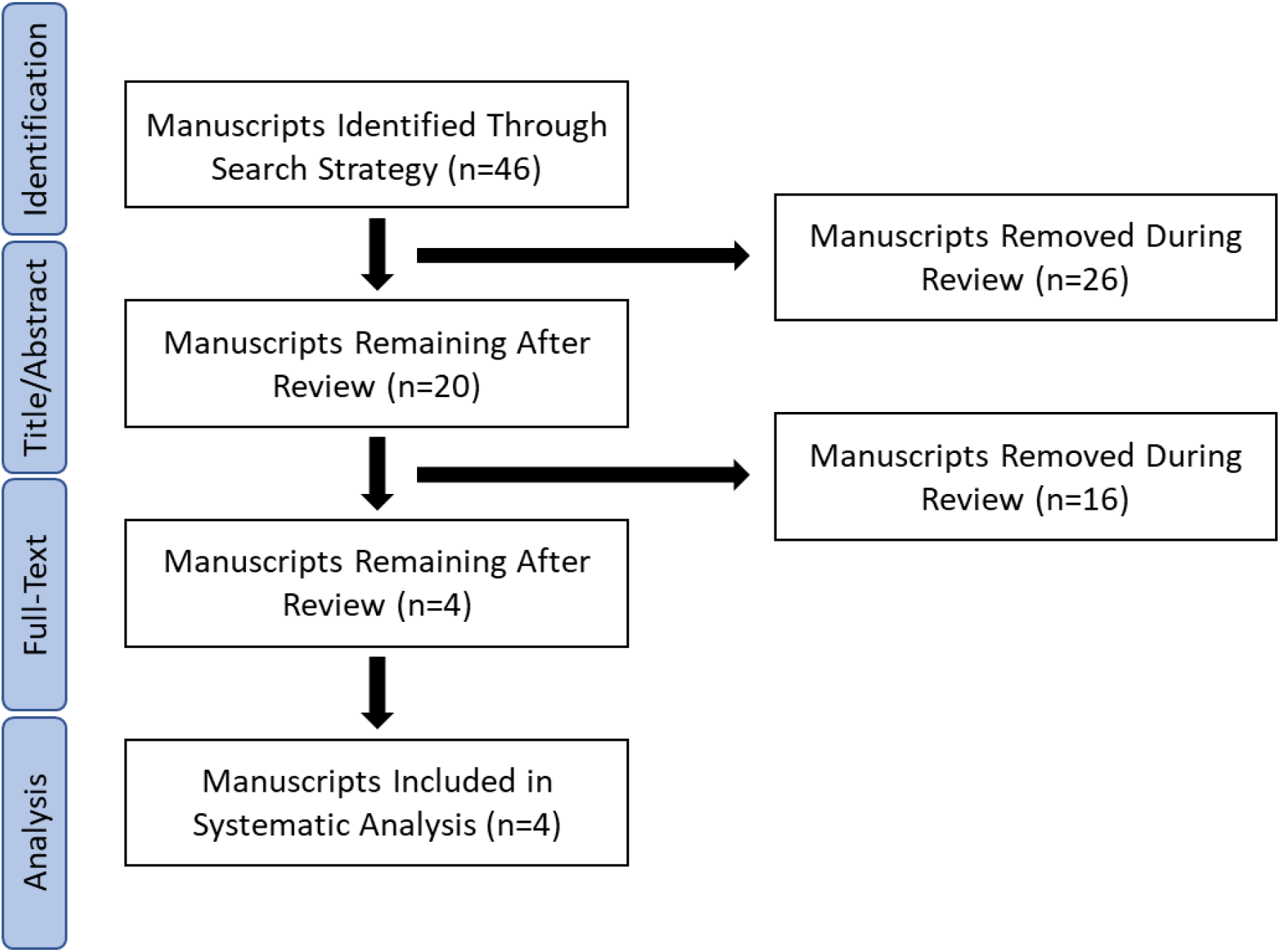
PRISMA flow chat. The flow chart in this figure depicts the methodology for the systematic literature review that was performed using the PubMed, EMBASE, and MedLine Plus databases. The title and abstract review was the first screening phase which determined 26 manuscripts to be irrelevant. The full-text review was the second screening phase which determined 16 manuscripts to be irrelevant. The final phase of the review was a systematic analysis. All decisions in the review phases were agreed upon by at least two reviewers.

First, the research team performed an initial screening of the title and abstract of each manuscript to determine eligibility. Two researchers independently assessed each candidate manuscript. Manuscripts that received discordant votes were discussed among the research team until consensus was reached. We then performed a full-text review of the remaining manuscripts. We again discussed and reached consensus on the manuscripts that received discordant votes. The relevant manuscripts identified at this stage proceeded to the final systematic analysis.

Only those works that described guidelines for public reporting during outbreaks were determined to be eligible for inclusion in the final analysis. Works presenting personal opinions and guidelines for non-infectious diseases, review articles including systematic or narrative reviews, analyses of outbreak data that lack recommendations on public reporting, duplicate studies, and studies discussing the delivery of clinical care were excluded from the study. This exclusion criteria ensured that only relevant guidelines to the specific topic of public reporting during outbreaks remained.

## Results

Our search strategy yielded 46 manuscripts. We excluded 26 manuscripts during the title and abstract review. Of those, 12 manuscripts received discrepant votes, in which the two reviewers did not agree on whether the manuscript should advance. Through discussion and deliberation, we reached a consensus in all cases.

The remaining 20 manuscripts advanced to full-text review. We excluded 16 of the 20 manuscripts due to irrelevance. Of those, 10 received discrepant votes. Again, we reached a consensus through further discussion and deliberation. Four manuscripts proceeded to the final analysis. Three manuscripts discuss results in local contexts, notably Iran, India, and the United States. The recommendations drawn from the manuscripts pertaining to Iran and India specifically are likely more broadly applicable to other contexts.^32,33^

The first manuscript of relevance to our systematic review, “Enhancing Situational Awareness to Prevent Infectious Disease Outbreaks from Becoming Catastrophic” by Lipsitch and Santillana, describes four key areas where situation reports can inform and assist policymakers in decision-making during an outbreak. Those areas are disease severity, epidemic size and geographic extent, transmissibility, and countermeasure availability, status and effectiveness.^34^ The authors recommend discussing sources of uncertainty for each information type, such as whether current surveillance systems have capacity to determine case counts to measure epidemic size and geographic extent.

The authors also highlight principles of high-quality data reporting that could be pertinent to the development of universal guidelines. The principles emphasize the conciseness, clarity, and value of information in decision-making. Subject matter expert assessments should accompany evidence gathered from multiple data sources, and situation reports should include curated visualizations and acknowledge uncertainty. These recommendations fall short of proposed or adopted consensus guidelines for public reporting. Nevertheless, they emphasize the importance of accurate and clear situation reporting for planning and evidence-based decision-making during an outbreak, with relevant key areas that may inform the development of minimum information reporting standards.

The second manuscript, “Coronavirus disease 2019 (COVID-19) surveillance system: Development of COVID-19 minimum data set and interoperable reporting framework” by Shanbehzadeh et al., describes the standardization of a minimum COVID-19 data set and data entry process to enable interoperability across clinical and public health information systems in Iran.^32^ The proposed Minimum Data Set (MDS) includes administrative and clinical data categories, each finalized through Delphi surveys. Shanbehzadeh et al. do not explicitly aim to develop guidelines for public reporting during infectious disease outbreaks. Instead, they seek to integrate the information life cycle that underpins the COVID-19 surveillance system. Nevertheless, Shanbehzadeh et al.’s suggested MDS data categories offer potentially relevant reporting items. The administrative data category includes admission, demographic information, and report ID. The clinical data category comprises clinical presentation, CT results, discharge outcome, exposure to causal factors, laboratory findings, physical examination, signs and symptoms, and treatment plan. In addition to establishing MDS data categories, Shanbehzadeh et al. identified unreliable data entry and poor uptake as limiting the quality of surveillance data. They recommend developing and incorporating user-friendly, compulsory data fields rather than free-text writing. These recommendations emphasize efficiency and consistency and could refine the design of consensus guidelines.

The third manuscript, “Variation in COVID-19 Data Reporting Across India: 6 Months into the Pandemic” by Vasudevan et al., offers a comprehensive evaluation of inter-state variations in data reporting quality for COVID-19 cases across India between 12 July and 25 July 2020.^33^ The authors examined the quality of state data for confirmed, deceased, recovered, quarantined and Intensive Care Unit (ICU) cases according to four criteria: availability of data (daily and cumulative counts), accessibility of data, granularity (covering age, gender, co-morbidities, and districts), and data privacy. Vasudevan et al. selected these scoring metrics with the understanding that essential public health messages should reach audiences beyond the scientific community. The metrics could help refine public reporting standards that reflect the public’s needs during infectious disease outbreaks. Finding an apparent strong variability in COVID-19 data reporting, Vasudevan et al. seek to assist states in improving the quality of their reporting. They do not provide a formal assessment to develop, design, refine or establish consensus guidelines. However, the authors highlight the difficulties of data aggregation in the absence of a unified reporting framework and its consequent negative effects on coordinated and effective nationwide responses to the pandemic.

The fourth manuscript, “Evaluating Completeness of Foodborne Outbreak Reporting in the United States, 1998-2019” by Zhang et al., describes the completeness and frequency of outbreak reports by seasonal and annual trends and by pathogen across the Centers for Disease Control and Prevention’s electronic Foodborne Outbreak Reporting System and the National Outbreak Reporting System.^35^ Zhang et al.’s efforts, while not specifically aimed at establishing minimum information reporting standards and expectations by governments during an outbreak, offer potentially relevant recommendations. The authors first recommend developing a standard operating procedure to help streamline the data cleaning process and to determine which reporting variables are necessary and interrelated in outbreak reports. They also recommend removing reporting variables consistently low in completeness and publicly reporting the reasons for incompleteness. Finally, they suggest periodically auditing data reporting procedures, including the quality of data reporting practices at local levels.

## Conclusion

The four manuscripts identified in this study highlight important features of data reporting quality and content for decision-makers and wider audiences during an outbreak. Together, they show the discretion afforded to local public health authorities in determining when, where, how, and what data to report on outbreaks. This flexibility results in discrepancies in the consistency and quality of data reporting, which can hamper outbreak detection and response efforts. The absence of universal minimum information reporting standards and expectations by governments is apparent and clearly outlines both the opportunity and need for their development.

Examining the recommendations from each manuscript provides a starting point in determining what a set of minimum standards and expectations for outbreak reporting can look like. The ideal outbreak report could contain any of the following data points: disease severity, size and geographic extent, daily and total case count, demographics, transmissibility, signs and symptoms, probable disease transmission and exposure pathways, countermeasure status, and sources of uncertainty.^32–35^ The report should be clear and digestible, aided by carefully curated visualizations and subject matter expertise.^34^ Finally, the ideal outbreak report should provide at least the minimum data points needed to inform major outbreak response decisions.^32–35^

This systematic literature review surveyed previous efforts to achieve universal minimum standards for outbreak data reporting. We propose developing and promoting consensus guidelines for the public reporting of outbreak data as a next step and, consequently, have launched the Enabling Accountability for Effective Outbreak Reporting Best Practices in Transparency (ORBIT) project. Through ORBIT, we aim to establish minimum standards and expectations for information reporting during an outbreak and to promote their widespread adoption. A consensus panel of experts in epidemiology, outbreak response, and emergency operations across academia, government and non-governmental organizations will generate the complete list of reporting items. The systematic literature presented in this paper substantiates the need for ORBIT guidelines and serves as a basis for relevant data points. We expect to produce results from the ORBIT guideline development efforts in 2024.

## Data Availability

All data produced in the present work are contained in the manuscript

## Acknowledgments

The research team would like to acknowledge the Open Philanthropy Project for providing support for this work. The funders had no input or oversight over the conduct of this work.

The authors also acknowledge Erin Fink for her contribution to the study conceptualization, methodology, data collection and formal analysis. The authors also acknowledge Noelle Huhn for her contributions to the study conceptualization and methodology.

